# Strategies for using antigen rapid diagnostic tests to reduce transmission of SARS-CoV-2 in low- and middle-income countries: a mathematical modelling study applied to Zambia

**DOI:** 10.1101/2022.06.16.22276516

**Authors:** Alvin X. Han, Sarah Girdwood, Shaukat Khan, Jilian A. Sacks, Amy Toporowski, Naushin Huq, Emma Hannay, Colin A. Russell, Brooke E. Nichols

## Abstract

**Background:** Increasing the availability of antigen rapid diagnostic tests (Ag-RDTs) in low- and middle-income countries (LMICs) is key to alleviating global SARS-CoV-2 testing inequity (median testing rate in December 2021-March 2022 when the Omicron variant was spreading in multiple countries; high-income countries=600 tests/100,000 people/day; LMICs=14 tests/ 100,000 people/day). However, target testing levels and effectiveness of asymptomatic community screening to impact SARS-CoV-2 transmission in LMICs are unclear.

**Methods:** We used PATAT, an LMIC-focused agent-based model to simulate COVID-19 epidemics, varying the amount of Ag-RDTs available for symptomatic testing at healthcare facilities and asymptomatic community testing in different social settings. We assumed that testing was a function of access to healthcare facilities and availability of Ag-RDTs. We explicitly modelled symptomatic testing demand from non-SARS-CoV-2 infected individuals and measured impact based on the number of infections averted due to test-and-isolate.

**Results:** Testing symptomatic individuals yields greater benefits than any asymptomatic community testing strategy until most symptomatic individuals who sought testing have been tested.

Meeting symptomatic testing demand likely requires at least 200-400 tests/100,000 people/day on average as symptomatic testing demand is highly influenced by non-SARS-CoV-2 infected individuals. After symptomatic testing demand is satisfied, excess tests to proactively screen for asymptomatic infections among household members yields the largest additional infections averted.

**Conclusions:** Testing strategies aimed at reducing transmission should prioritize symptomatic testing and incentivizing test-positive individuals to adhere to isolation to maximize effectiveness.

## Introduction

Since the emergence of SARS-CoV-2 in 2019, the COVID-19 pandemic has resulted in over 500 million confirmed cases and 6 million deaths worldwide as of May 2022.[1] While vaccination is the key medical intervention to mitigate the pandemic, SARS-CoV-2 testing remains an important public health tool for case identification and transmission reduction. Testing is especially important in many low- and middle-income countries (LMICs) that continue to struggle with gaining equitable access to global vaccine supplies.[2] Testing is also the backbone of surveillance systems to monitor the emergence of novel variants of concern (VOCs)[3] that may escape immunity acquired from previous infections and vaccination.[4]

At the same time, the global imbalance in SARS-CoV-2 testing rates is substantial.[5] Between December 2021 and March 2022 when the Omicron (BA.1) VOC was spreading in multiple countries, the median testing rate in LMICs was 14 (IQR = 7-41) tests per 100,000 persons per day (/100k/day), whereas HICs tested >43 times more, with a median rate of 603 (IQR = 317-1181) tests/100k/day.[6] Limited testing has likely led to substantial underestimation of SARS-CoV-2 prevalence and COVID-19 attributable mortality in LMICs.[7] The diagnostics pillar of the Global Access to COVID-19 Tools (ACT) Accelerator, co-convened by FIND and the Global Fund in partnership with the World Health Organization to enhance access to COVID-19 tests and sequencing, has set a minimum testing rate of 100 tests/100k/day.[8] This minimum testing rate is thought to be a “critical threshold to facilitate effective public health interventions”.[8] Additionally, asymptomatic testing in a community setting (i.e. community testing) was identified by this initiative as a crucial step for LMICs to close in on the global equity gap.[8]

Real-time reverse transcription polymerase chain reaction (PCR) tests remain the gold standard for COVID-19 diagnostic testing, as they are the most sensitive testing method.[9] However, PCR-based testing can be plagued by long turnaround times and necessitate relatively costly laboratory infrastructures, robust sample transport networks and well-trained personnel that are lacking in many low-resourced settings.[10] Furthermore, RNA can still be detected even after infectiousness has declined, rendering PCR tests imperfect for determining the infectious potential of an infected person.[11] While the sensitivity of antigen rapid diagnostic tests (Ag-RDTs) is lower than PCR(>80%)[12], Ag-RDTs are cheaper, capable of producing results in under 30 minutes and can be performed easily at point-of-care.[13] As such, when used in a timely fashion, Ag-RDTs can identify potentially infectious people more quickly. Ag-RDTs offer a practical alternative diagnostic tool to enable massive scale up of testing in all countries. In resource-limited settings, Ag-RDTs could potentially reduce the testing equity gap between HICs and LMICs.[5]

To date, there is no robust evidence-base on how scaling-up Ag-RDTs to 100 tests/100k/day would impact community transmissions. There is also a lack of information on the effectiveness of community testing programs when used to complement symptomatic testing under the constraints of limited test availability. Furthermore, while there have been studies investigating the impact of comprehensive test-and-trace programs on transmission reductions,[14,15], it is less clear to what extent a test-and-isolation strategy (i.e. the only intervention is the required isolation of individuals with a positive diagnosis) would impact total infections. It is thus important to estimate the impact of test-and-isolation only since most low-resource settings did not implement resource-intensive contact tracing programs.[16]

In this study, we developed and used the Propelling Action for Testing and Treating (PATAT) simulation model, an agent-based modelling framework to investigate the impact of using Ag-RDTs for healthcare facility-based symptomatic testing. We considered testing programs both with and without additional asymptomatic testing programs in the community, using a population with demographic profiles, contact mixing patterns, and levels of public health resources akin to those in many LMICs. We used PATAT to interrogate how different Ag-RDT distribution availability and testing strategies, including the implementation of community testing in households, schools, formal workplaces and regular mass gatherings such as religious gatherings, could impact onward disease transmission. In turn, we aimed to identify key priorities and gaps that should be addressed when implementing mass testing programs using Ag-RDTs in low-resource settings.

## Methods

### The **P**ropelling **A**ction for **T**esting **A**nd **T**reating (PATAT) simulation model

PATAT first creates an age-structured population of individuals within contact networks of multi-generational households, schools, workplaces, religious gatherings (i.e. regular mass gatherings) and random community with the given demographic data here based on archetypal LMIC estimates (Figure 1). The simulation starts with a user-defined proportion of individuals infected with SARS-CoV-2. Given that viral loads of an infected individual at the time of testing affect Ag-RDT sensitivity,[12] PATAT randomly draws a within-host viral load trajectory over the course of each individual’s infection from known distribution of trajectories[17,18] using previously developed methods.[19] Given conflicting evidence,[20] similar viral load trajectories were drawn for both asymptomatic and symptomatic infected individuals.

**Figure 1:**
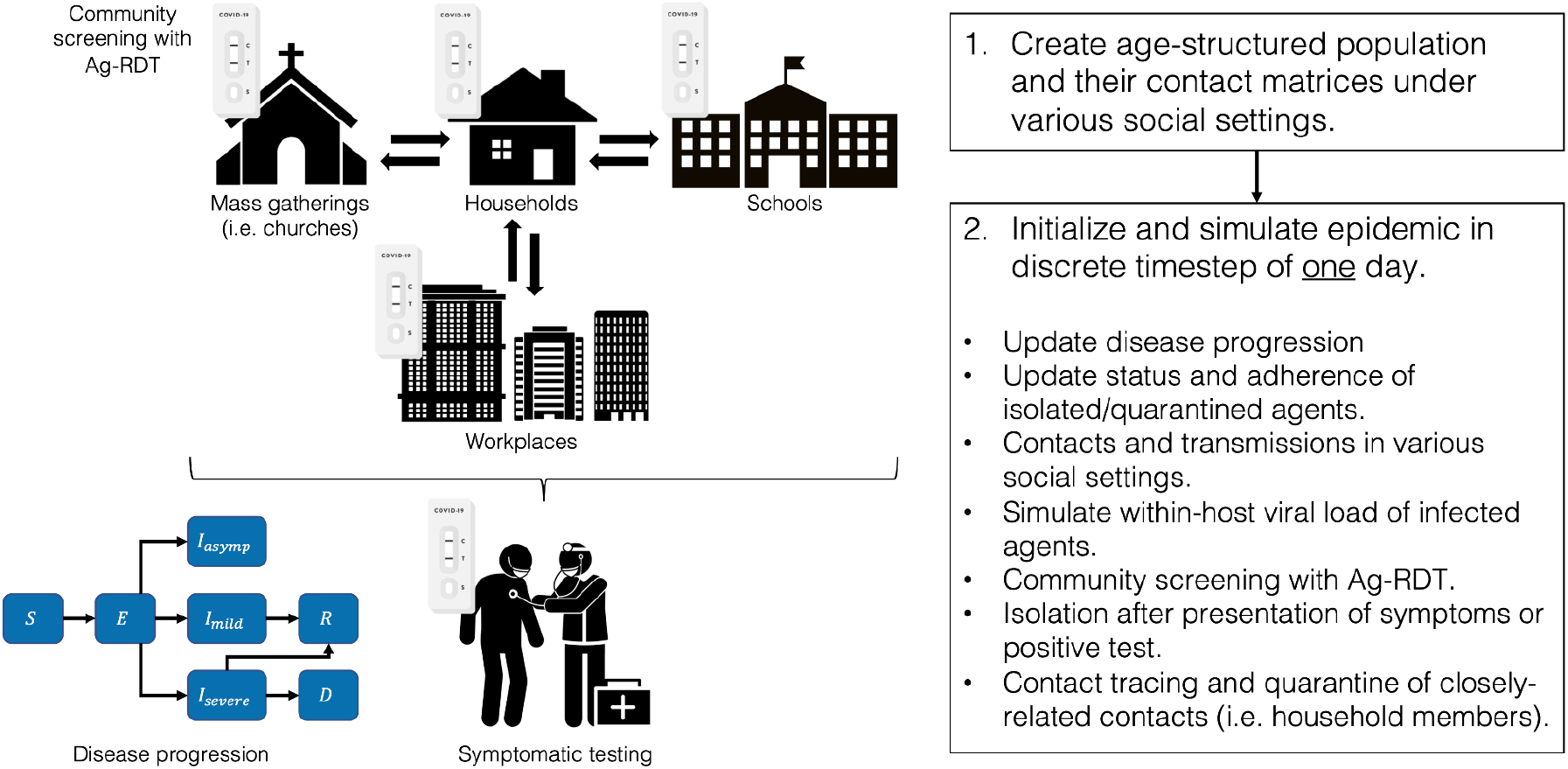
Schematic of <u>P</u>ropelling <u>A</u>ction for <u>T</u>esting <u>A</u>nd <u>T</u>reating (PATAT) simulation model.

The simulation computes transmission events across different contact networks each day and updates the disease progression of infected individuals based on the Susceptible-Exposed-Infected-Recovered-Death (SEIRD) epidemic model, stratifying them based on symptom presentation (asymptomatic, mild or severe). Symptomatic individuals may seek symptomatic testing at clinics after symptom onset. Given that most LMICs are currently testing at rates far below 100 tests/100k/day to the extent that only a small proportion of COVID-19 positive deaths were identified in life (e.g. <10% in Zambia),[21] we assumed that all clinic-provided testing demand by mild symptomatic individuals are satisfied by Ag-RDTs while PCR tests are restricted for testing severe patients only. Positively-tested individuals may go into isolation and their household members may also be quarantined.

### Simulation variables

We assumed a populations size of 1,000,000 individuals, creating contact networks and healthcare facilities based on demographic data collected from Zambia.[22] We initialized each simulation with 1% of the population being infected by SARS-CoV-2 and ran the model over a 90-day period. We permutated a range of *R*_*e*_ values (i.e. 0.9, 1.1, 1.2, 1.5, 2.0, 2.5 and 3.0) against varying Ag-RDT stock availability (i.e. 100, 200-1,000 (in 200 increments), 1,000-5,000 (in 1,000 increments) tests per 100,000 persons per day). Various test distribution strategies were simulated: (1) 85% of weekly allocated tests were used for routine asymptomatic community testing at a social setting and the remaining used for symptomatic testing at healthcare facilities; (2) all weekly allocated test stocks are distributed to healthcare facilities for symptomatic testing only with no asymptomatic community testing and (3) weekly allocated tests are used for symptomatic testing at healthcare facilities first before any remaining tests at the end of the week are used for asymptomatic community testing in the next week. As a baseline, we simulated a set of runs using the same range of *R*_*e*_ with no testing at all.

To determine impact of testing on reducing infections, we assumed that the only public health intervention measure is by test-and-isolate of positive-tested individuals. We also performed a separate set of simulations that require a same-day quarantine of asymptomatic household members of positively-tested individuals. This distinction is important because quarantine should change contact patterns of more individuals per positive test, thereby increasing test utility. We did not consider the quarantine of close contacts outside of household members as contact tracing programs are often resource intensive and discontinued in most countries.

### Distribution of routine asymptomatic community test

Due to their fixed nature and potential accessibility, routine asymptomatic community testing may be implemented in households, schools, formal workplaces or religious gatherings. Community tests stocks may be distributed in each setting in two ways: (1) even distribution to as many entities as possible once per week (e.g. if we have 10 tests available for 10 households per week, then one member of each household would be tested); (2) concentrated distribution to test all individuals in selected entities twice a week who will continue to get tested throughout the epidemic (e.g. if we have 10 tests available for 10 households per week but only one housing of 5 members, then all 10 tests will be distributed to this selected household of 5 for testing on Monday and Thursday of every week).

### Healthcare provided symptomatic testing demand

Symptomatic testing demand estimation is particularly challenging for SARS-CoV-2 because COVID-19 symptoms overlaps with other respiratory infections, thus increasing testing demand. We assumed that symptomatic individuals would seek testing at clinics based on a probability distribution that inversely correlate with the distance between their homes and the nearest clinic[23] (Table S1). We assumed that the time delay between testing and symptom onset follows lognormal distribution of with mean of one day and standard deviation of 0.5 day. Additionally, we simulated daily demand of clinic tests from individuals that were not infected by SARS-CoV-2 but sought symptomatic testing as they presented with COVID-19-like symptoms. This non-COVID-19 related demand was estimated by assuming a 10% test positivity rate at the start as well as end of an epidemic curve and 20% test positivity rate at the peak, linearly interpolating the demand for periods between these time points (Figure 2). These assumptions are based on observed test positivity rates in multiple countries experiencing infection waves during the second half of 2021.[24] If there are limited clinic test stocks for the day, the available tests are randomly distributed among symptomatic SARS-CoV-2-infected patients and those seeking tests for non-COVID-19 related reasons. We assumed that any individual who failed to receive a test due to test shortage would not seek clinic-provided testing again for the rest of their infection. If these individuals had previously decided to self-isolate upon presenting symptoms, they may continue to do so (see Supplementary Data). Otherwise, we assumed that they would continue to mix with the community. We also assumed that SARS-CoV-2 infected individuals who were tested but received a false negative result continue mixing with the community. In turn, any false positive tested individual would then go into isolation.

**Figure 2:**
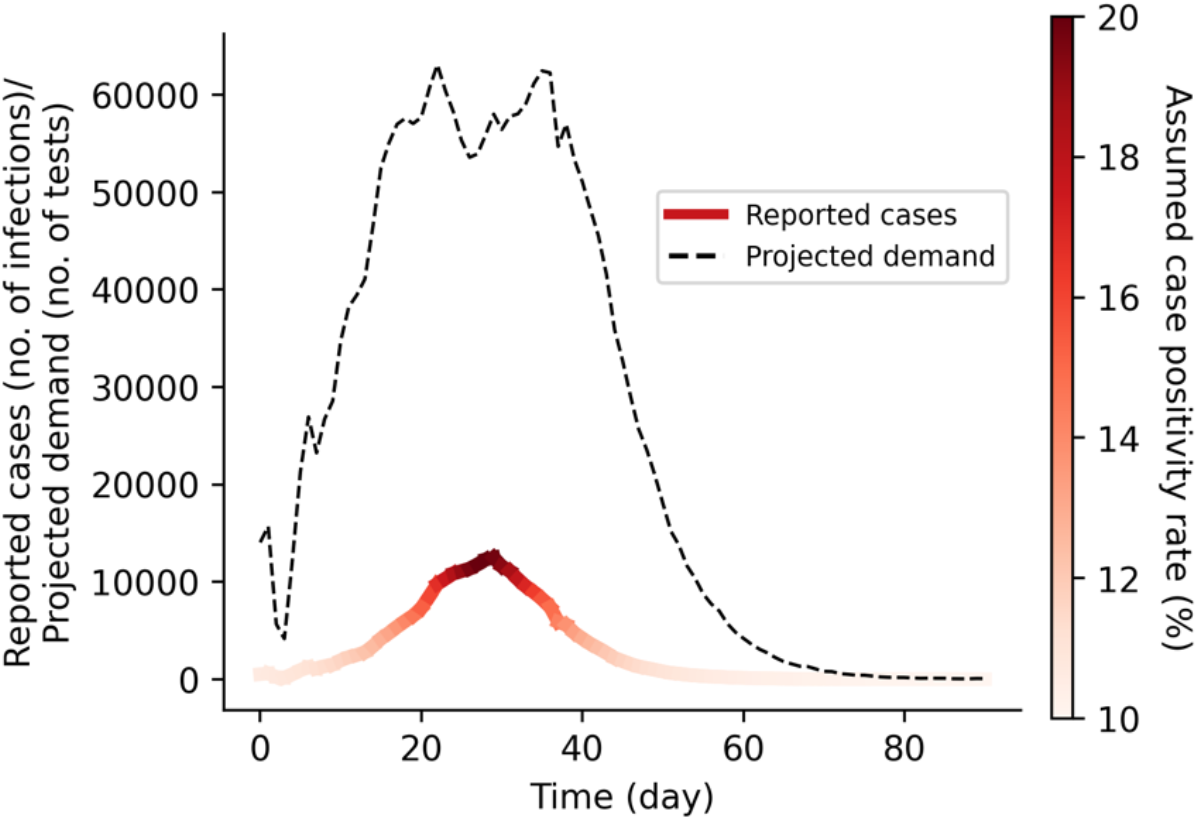
Projected symptomatic testing demand based on assumed case positivity rate. This projected demand includes both SARS-CoV-2 infected persons who were tested and reported as well as those who seek symptomatic testing for other reasons (e.g. individuals presenting COVID-19-like symptoms but were not infected with SARS-CoV-2).

All key parameters are tabulated in Table S1 and full details of PATAT are described in the Supplementary Data. The PATAT model source code is available at https://github.com/AMC-LAEB/PATAT-sim.

## Results

### Healthcare provided symptomatic testing demand should be fulfilled first

We first investigated if routine asymptomatic community testing programs could substantially reduce SARS-CoV-2 transmissions in the population. We compared scenarios where either all Ag-RDT stocks were used only for symptomatic testing (with no community-based testing of asymptomatic populations), or that most tests were used for community testing and only 15% of weekly available stocks were allocated for symptomatic testing. The proportion of infections averted under each test distribution strategy was computed as a measure of impact. Regardless of community testing distribution strategy or *R*_*e*_, we found that setting aside large proportions of Ag-RDTs for community testing led to lower proportion of infections averted than if all tests were solely used for symptomatic testing (Figure 3). A far greater number of tests is needed under the community testing scenarios relative to the only-symptomatic one to result in an equal or larger proportion of infections averted. This conclusion remains the same when household members of all positively tested individuals were quarantined (Figure S1).

**Figure 3:**
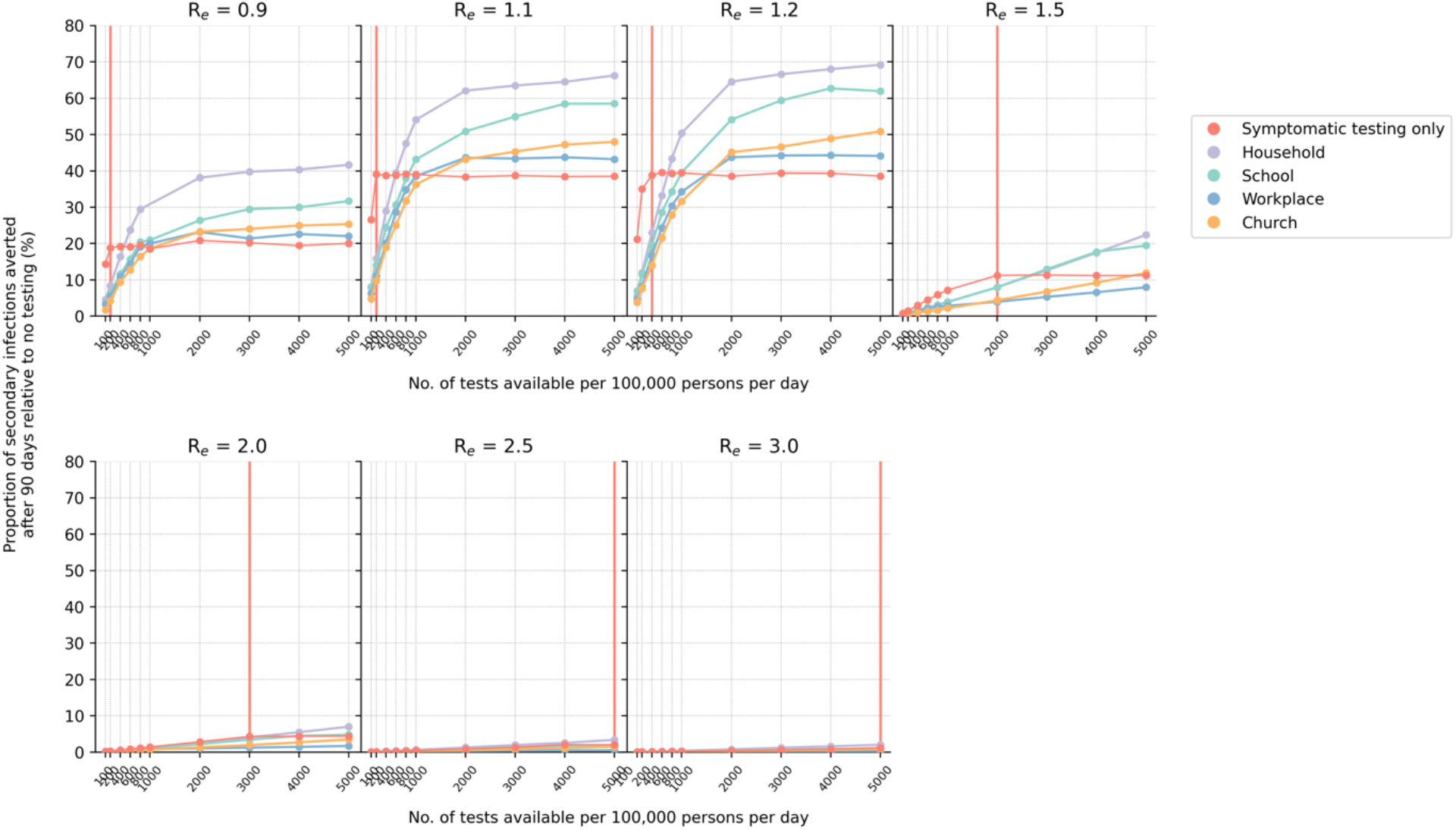
Impact of either using all available Ag-RDT for symptomatic testing or a majority of them (85%) for community testing in various settings (even distribution only; <u>without</u> quarantine of household members). The proportion of secondary infections averted after 90 days relative to the no testing baseline for different number of tests available per 100,000 persons per day and assumed *R*_*e*_ value is plotted for each test distribution strategy. The vertical red line denotes the number of tests required to saturate symptomatic testing demand (Figure 4).

### Number of tests needed to saturate healthcare provided symptomatic testing demand

Given the importance of saturating symptomatic testing demand, we then estimated the number of tests needed to saturate symptomatic testing demand under different *R*_*e*_ (Figure 3 and 4A). Besides symptomatic SARS-CoV-2 infected individuals who sought testing, our simulations factored in that 80-90% of symptomatic test stocks were used by individuals who were not infected by SARS-CoV-2 by assuming that test positivity rate ranges at 10%-20% over the course of the infection wave. Under these assumptions, even when *R*_*e*_ ≤ 1.2 (Figure 4B), at least 200-400 tests/100k/day was needed to ensure all test-seeking individuals were tested. If *R*_*e*_ ≥ 1.5, at least 10 times more tests, in the range of 2,000-5,000 tests/100k/day, was needed to satisfy all symptomatic testing demand. These conclusions were similar when household members of positive-tested individuals were quarantined (Figure S2).

**Figure 4:**
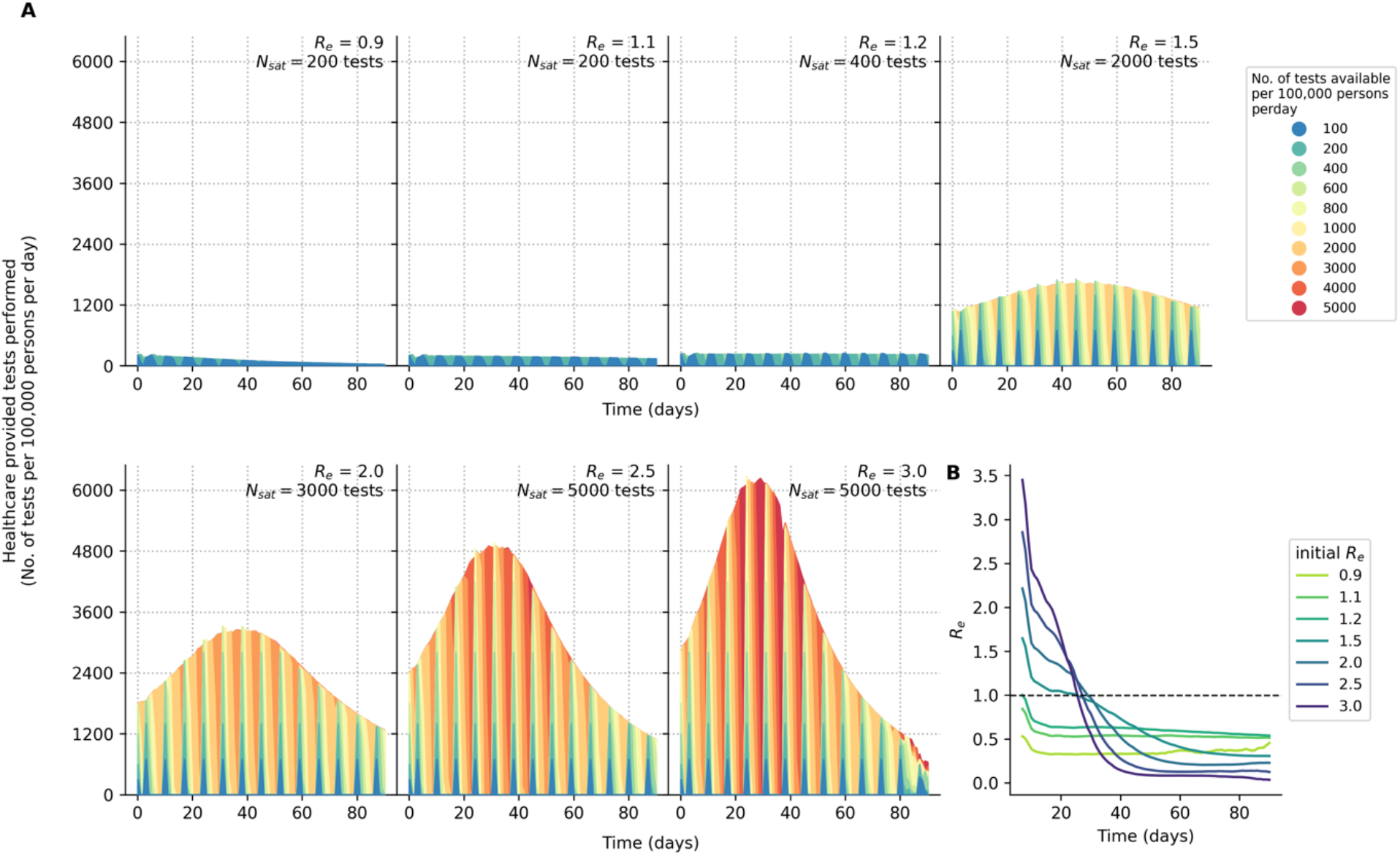
Symptomatic testing demand during an epidemic (<u>without</u> quarantine of household members). (**A**) Number of symptomatic tests performed per 100,000 persons per day over time for different *R*_*e*_. Each differently colored shaded curve denotes a different number of tests available per 100,000 persons per day. We assumed that all healthcare facilities in the community will have new stocks of one week’s worth of Ag-RDTs every Monday. The symptomatic testing demand include both symptomatic SARS-CoV-2 infected individuals who seek testing at healthcare facilities and those who seek symptomatic testing for other reasons based on assumed case positivity rates (see Methods). The area between the curve plotting the testing rate needed to saturate symptomatic testing demand (*N*_*sat*_) and the curve for testing rate < *N*_*sat*_ is the amount of symptomatic testing shortage accumulated over time between those two testing rates. (**B**) 7-day moving average of time-varying effective reproduction number (*R*_*e*_) over simulated epidemic period (90 days) assuming that testing demand is fully satisfied.

### Marginal impact of symptomatic testing prior to saturating demand

We linearly regressed the number of infections averted against test availability to compute the number of additional infections averted with increasing Ag RDT availability, before saturating symptomatic testing demand (Figures 5A-B). Assuming only symptomatic cases that test positive isolate, the largest marginal benefit of increasing Ag-RDT availability for symptomatic testing prior to demand saturation is achieved when *R*_*e*_ = 1.1-1.2, with close to 20,000 additional infections averted for every increase of 100 more Ag-RDTs available for symptomatic testing (Figure 5B; Table 1). When operating at tests availability that meet all symptomatic testing demand, the greatest impact of test-and-isolate is also achieved when *R*_*e*_ = 1.1-1.2 with ∼40% of total infections averted (Figure 5A).

**Figure 5:**
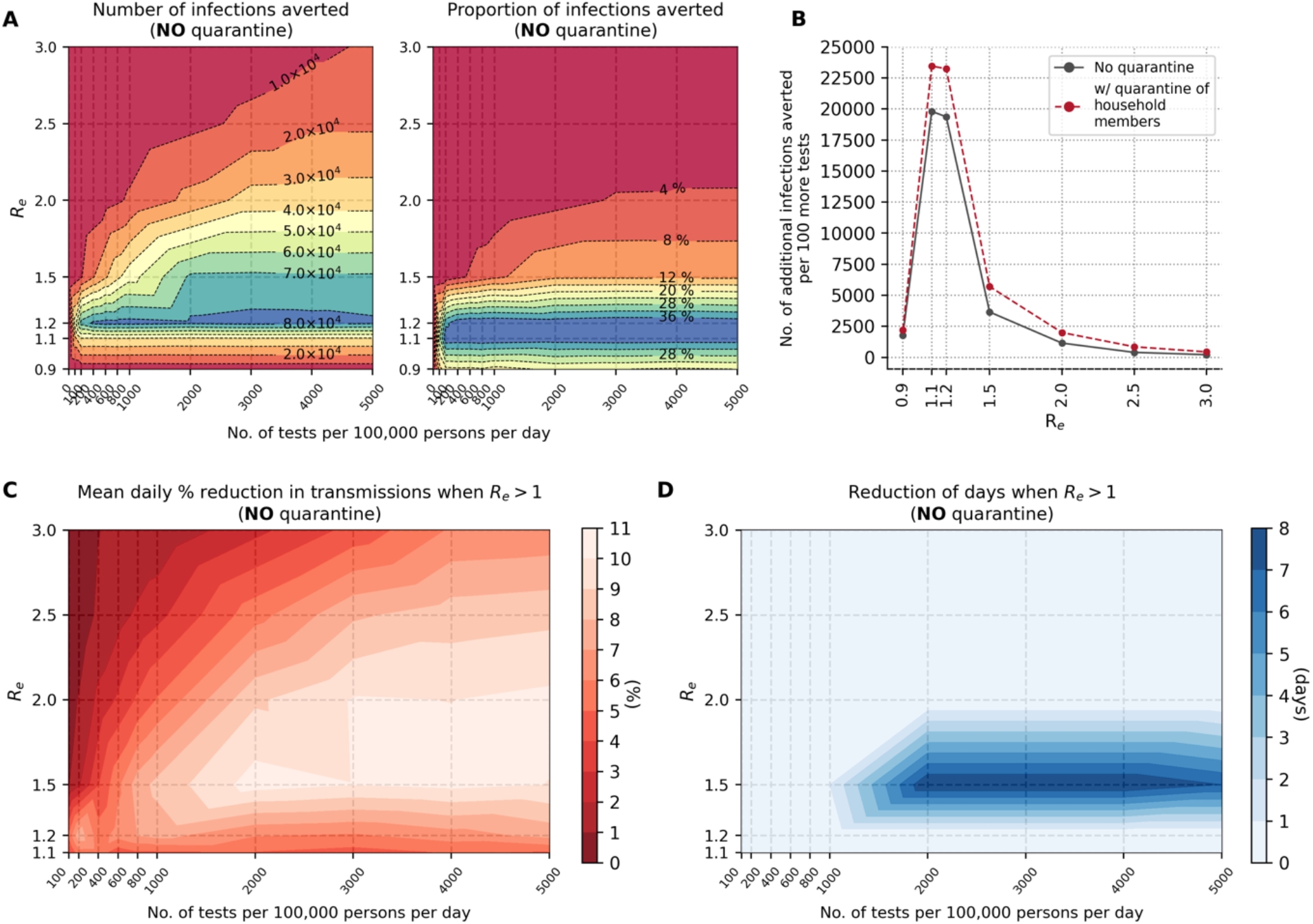
Marginal impact of symptomatic testing prior to saturating demand (<u>without</u> quarantine of household members). (**A**) Contour plots depicting infections averted relative to the no testing baseline for simulations with different *R*_*e*_values and varying number of Ag-RDTs availability. Number of infections averted relative to no testing baseline after 90 days (left panel); Proportion of secondary infections averted relative to no testing baseline after 90 days (right panel). (**B**) Number of additional infections averted for every 100 more Ag-RDTs available prior to saturating symptomatic testing demand for different *R*_*e*_values. Dashed red line shows marginal benefit with quarantine of household members while solid black line depicts that without quarantine. (**C**) Mean daily percentage reduction in transmissions while time-varying *R*_*e*_ of simulated epidemic is still > 1 for different initial *R*_*e*_values and varying number of Ag-RDTs available for symptomatic testing only. (**D**) Reduction in number of days when time-varying *R*_*e*_ of simulated epidemic is > 1 for different initial *R*_*e*_values and varying number of Ag-RDTs available for symptomatic testing only.

**Table 1:** Number of additional infections averted for every 100 more Ag-RDTs available prior to saturating symptomatic testing demand for different *R*_0_values.

| w/ quarantine of household members | $R_e$ | No. of additional infections averted per 100 more tests |
| --- | --- | --- |
| No | 0.9 | 1,772 |
|  | 1.1 | 19,807 |
|  | 1.2 | 19,372 |
|  | 1.5 | 3,655 |
|  | 2.0 | 1,149 |
|  | 2.5 | 401 |
|  | 3.0 | 216 |
| Yes | 0.9 | 2,205 |
|  | 1.1 | 23,444 |
|  | 1.2 | 23,250 |
|  | 1.5 | 5,702 |
|  | 2.0 | 1,999 |
|  | 2.5 | 853 |
|  | 3.0 | 441 |

Both the marginal benefit and the maximum infection reduction at demand saturation, however, diminish exponentially with increasing values of *R*_*e*_ (Figure 5A-B and Table 1). Nonetheless, there are other impacts that could be gained from performing more symptomatic testing at values of *R*_*e*_ > 1.2. For instance, for *R*_*e*_ values between 1.5 and 2.0 without quarantining household members, it is possible to reduce daily transmissions by up to 11% with increasing levels of test availability during the growth phase of the epidemic (*R*_*e*_ > 1; Figure 5C). Additionally, when *R*_*e*_∼1.5 and test availability is in the range of 2,000 tests/100K/day or more, it is possible to shorten the duration of the epidemic’s growth phase (and in turn, the epidemic itself) by about one week (Figure 5D).

The marginal benefit of symptomatic testing can be further augmented if asymptomatic household members of positively-tested individuals quarantine as well (Figure S3). However, depending on *R*_*e*_ and level of test availability, the percentage of infections averted only improved modestly by 2-10%. As we assumed that individuals would isolate and quarantine in their own homes, infectious individuals in isolation may infect healthy household members in quarantine with them.

### A symptomatic-testing-first strategy to community testing

Given the importance of symptomatic testing, we then simulated an alternate community testing strategy that prioritizes saturating symptomatic testing demand first every week. If there were leftover tests from clinics in the previous week, they were used for community testing in the following week. We also investigated two ways in which community tests were either evenly and randomly distributed in the social setting to as many individuals as possible or concentrate the available tests to a fixed number of persons throughout the epidemic period.

Even under this symptomatic-testing-first approach, other than households, community testing in almost all social settings only yields greater reduction in infections when test availability is higher than what is needed to saturate symptomatic testing needs (Figure 6). Overall, household community testing yielded the greatest reduction in transmissions for all simulated *R*_*e*_ values, followed by schools if *R*_*e*_ ≤ 1.5. Community testing in religious gatherings and formal workplaces only results in modest improvements over symptomatic testing. An even distribution of community tests tends to produce larger reduction in infections. The difference between even and concentrated community test distributions also increases with larger test availability. These results were similarly observed when household members of positively-tested individuals were quarantined (Figure S4).

**Figure 6:**
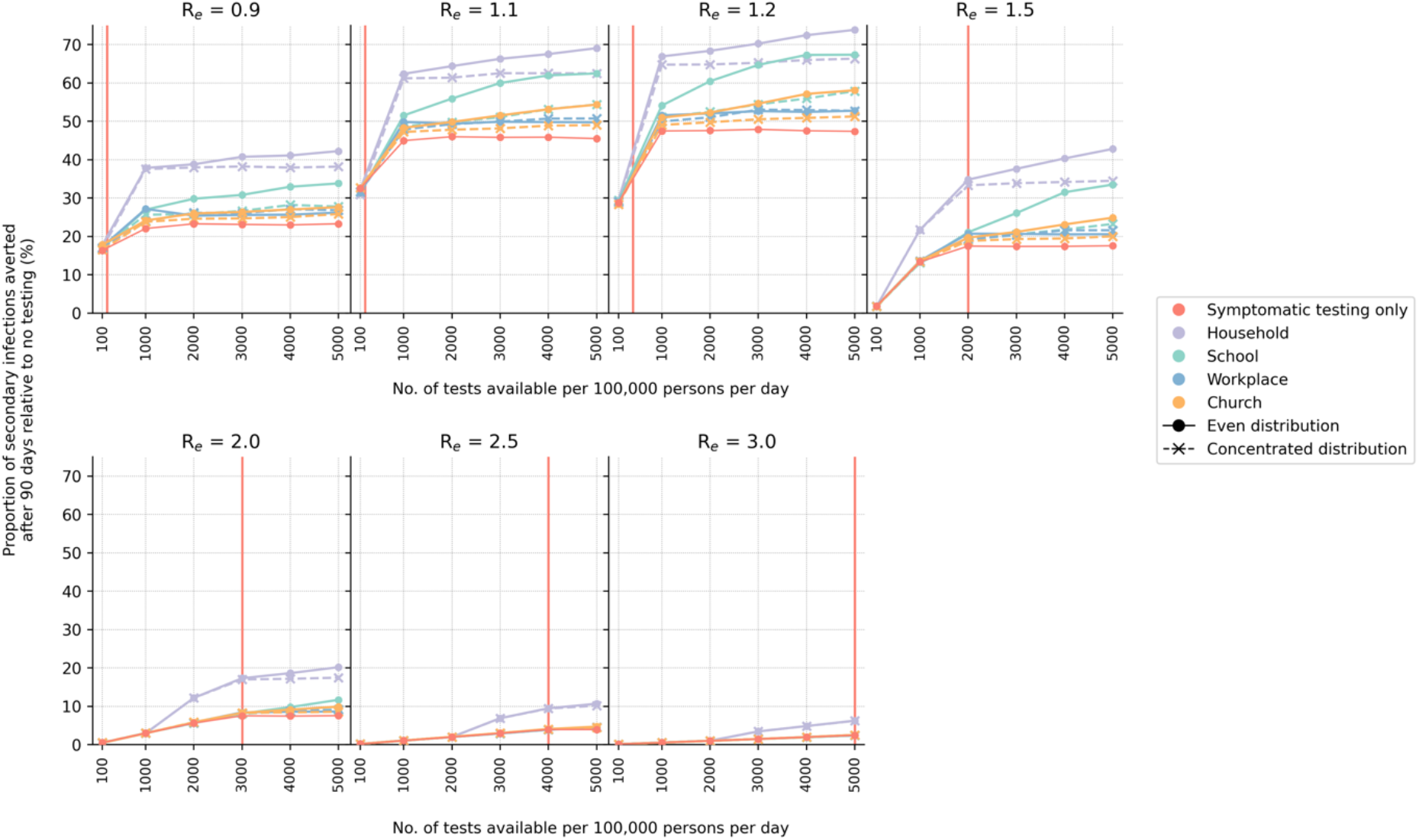
Symptomatic-testing-first strategy to community testing (<u>without</u> quarantine of household members). When community testing is performed under this strategy, the leftover tests from the previous week’s stock allocated for symptomatic testing are used for community testing in various setting in the current week. Two different types of community test distributions approaches (even or concentrated; see Methods) were simulated. The proportion of secondary infections averted after 90 days relative to the no testing baseline for different number of tests available per 100,000 persons per day and assumed *R*_*e*_ value is plotted for each test distribution strategy. The vertical red line denotes the number of tests required to saturate symptomatic testing demand.

### Routine community testing in households

In the symptomatic-testing-first approach, household community testing can achieve greater reduction in transmissions before saturating symptomatic testing demand but only at high levels of test availability (>1,000 tests/100k/day; Figure 6 and S4). There are several reasons why household community testing outperformed other settings. First, large multigenerational homes (mean household size = 5 people) were simulated to mirror what is often found in many LMICs. Second, population in LMICs tend to skew young (i.e. 48% of the population are expected to be ≤ 15 years in age).[22] Furthermore, overall employment rates are low (i.e. assumed 39% and 23% among men and women respectively)[22] and a large majority of employed individuals likely work in informal employment settings (i.e. assumed 64% and 76% among employed men and women respectively)[22] where test distribution is assumed to be difficult or infeasible. Third, dedicated isolation and quarantine facilities are likely rare in low-resource settings. Thus, positively-tested individuals and their close contacts could only isolate/quarantine themselves in their own homes. In turn, almost 60% of all infections observed in a typical simulation arose from transmissions in households. Random community transmissions aside, schools are then the second most common setting where transmissions occurred (∼14%) and workplaces, be if formal or informal, the least common (<3%) (Figure 7A).

**Figure 7:**
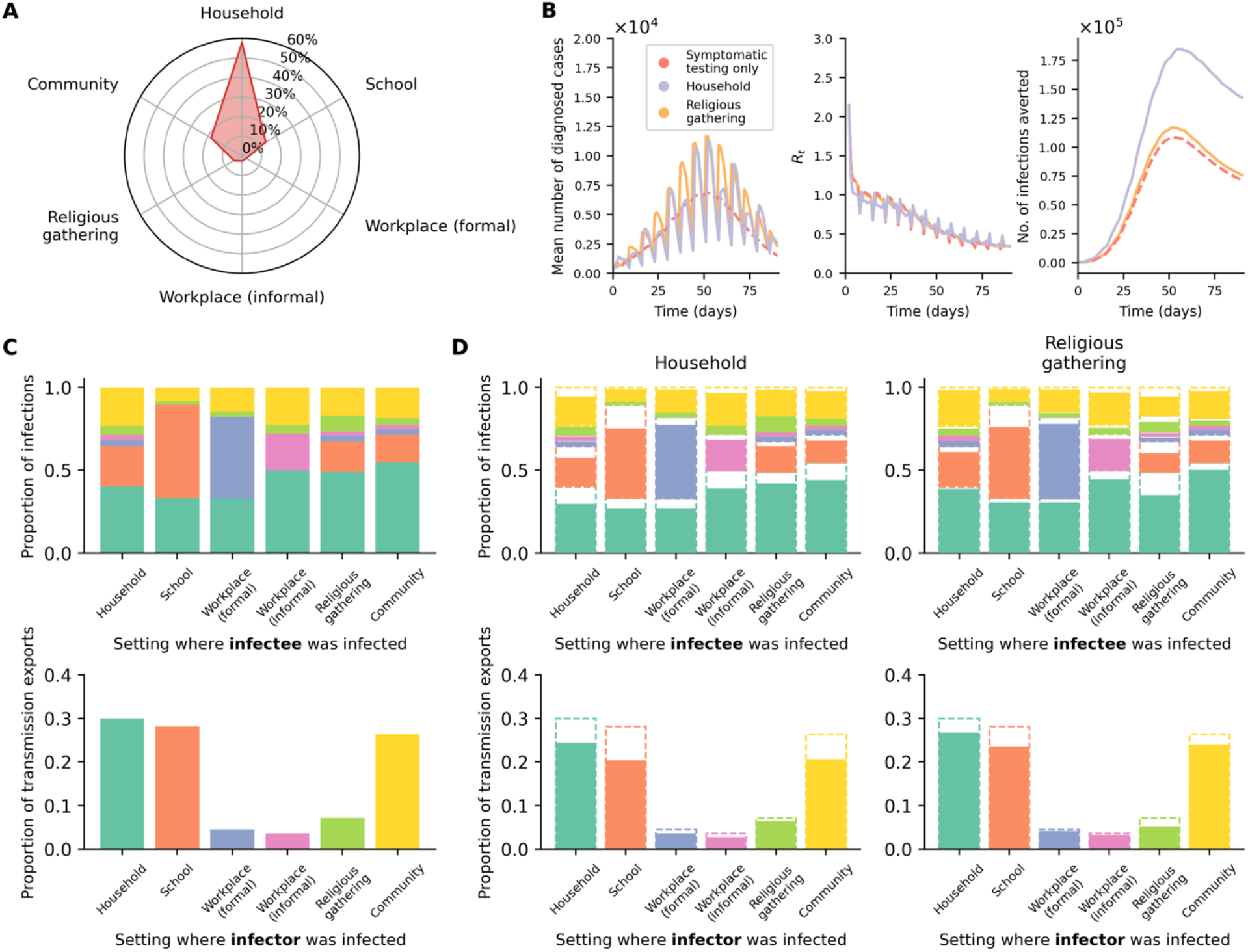
Routine community testing in households outperforms other settings. (**A**) Average breakdown of infections based on the social setting where transmissions occurred for the simulations presented in this work. (**B**) Results from simulations using different testing strategies where *R*_*e*_ = 1.5, no quarantine of household members of positively-tested individuals assumed, and Ag-RDT availability of 5,000 tests per 100,000 persons per day. Community testing (even distribution) was performed with a symptomatic-testing-first approach. The average total number of diagnosed cases (left), time-varying reproduction number (*R*_*t*_; middle) and number of infections averted (right) over the epidemic period are plotted. (**C, D**) Transmissions across distinct social settings. The top row of stacked plots shows the proportion of infections stratified by the source settings where infectors were infected for each sink setting where their infectees were infected. The stacked bars are colored by the source settings as per the bottom row of bar plots. The bottom row of bar plots shows the contribution of transmission exports into other settings (i.e. transmission events where the infectee were infected in a setting that is different from their infectors) from different source settings where the infectors were infected. (**C**) No testing baseline results from the example case as in (B). (**D**) Results from either implementing a symptomatic-testing-first community testing in households (left column) or religious gathering (right column). The dashed bar outlines are the no testing baseline results as in (C).

Interestingly, even though we assumed that 70% of all households regularly attended large religious gatherings weekly, they contributed to a limited proportion of total infections (∼5%) (Figure 7A). Yet, if we compare the results between household and religious gathering testing at levels of test availability large enough to satisfy symptomatic testing demand (e.g. *N* = 5,000), the total number of diagnosed cases over time is actually similar for both community testing strategies (Figure 7B). In fact, testing in religious gatherings yielded relatively larger number of cumulative diagnoses by the end of the epidemic but household testing suppressed *R*_*e*_ more during the growth phase of the epidemic, resulting in greater number of infections averted over time. This is because household testing not only reduces the already higher number of infections taking place in households, it also decreased transmissions between different distinct social settings (e.g. if an infector infected an infectee in the household setting but the infector was infected in school) (Figure 7C-D).

## Discussion

Community asymptomatic testing only achieves high levels of infection reduction after symptomatic testing demand has been saturated. However, the current minimum target of 100 tests/100k/day is unlikely to saturate symptomatic testing demand even with scenarios where *R*_*e*_ ≤ 1.2. Saturating symptomatic testing demand in realistic epidemic wave scenarios where *R*_*e*_ > 1.2 likely requires >1,000 tests/100k/day. This is because testing demand is largely shaped by non-SARS-CoV-2 infected individuals due to the overlap of SARS-CoV-2 infection symptoms with other respiratory tract infections. In other words, even before implementing any form of community testing, it is crucial to increase investments in testing capacities that meet symptomatic testing demand first. For instance, if Zambia had ∼10 million Ag-RDTs available through the first three months (i.e. ∼600 tests/100k/day over three months for 18 million people) of its first Omicron BA.1 wave (*R*_*e*_∼2.5) that were only used for symptomatic testing, ∼37,000 infections could likely be averted. However, if testing rate was at 100 tests/100k/day, the number of infections averted drops nearly 10-fold to ∼3700 cases averted on average despite only a 6-fold reduction in testing.

If *R*_*e*_ < 1.5, or can be reduced to that point through other public health interventions, increasing testing capacity from 100 tests/100k/day to 200-400 tests/100k/day provides the greatest proportional reduction in secondary transmissions. Furthermore, testing has the potential to be most effective at reducing transmission when *R*_*e*_ < 1.5. We would also obtain the greatest reduction in transmissions through increased testing volumes if *R*_*e*_∼1.0 (Table 1). As SARS-CoV-2 outbreaks can have *R*_*e*_ appreciably above 1.5, it is important to combine testing with other public health measures such as vaccination, physical distancing and masking so as to maximize impact of testing programs. It is also important to note that the utility of testing in averting infections is predicated on people changing and maintaining their behaviour to reduce contacts following a positive test.[25] Encouraging and incentivizing these changes of behaviour are essential for the effectiveness of any test-and-isolate program, particularly individuals of lower socioeconomic status[26] and communities in low-resource settings.[27]

As a corroboration of our results, we compared the weekly average testing rate[6] to the average *R*_*e*_ values estimated from COVID-19 case counts (https://github.com/epiforecasts/covid-rt-estimates)[28] of 134 countries between December 2021 and March 2022 when the Omicron BA.1 VOC spread rapidly across multiple countries (Figure 8). Although the demographic profiles differ between high-income countries (HICs) and LMICs, we found that some HICs were expectedly testing at rates that were sufficient or even higher than what was likely needed to saturate the symptomatic testing demand we had estimated for LMICs at similar epidemic intensity (i.e. *R*_*e*_ values). However, as Omicron (BA.1) cases surged, some HICs such as the United States, Germany and Australia were still reportedly facing test shortages.[29–31] Based on our results, these countries were indeed falling short of meeting symptomatic testing demand (Figure 8). Finally, if we assume that most HICs are testing at rates that sufficiently meet symptomatic testing demand, we found that most of them were testing >100 tests/100k/day.

**Figure 8:**
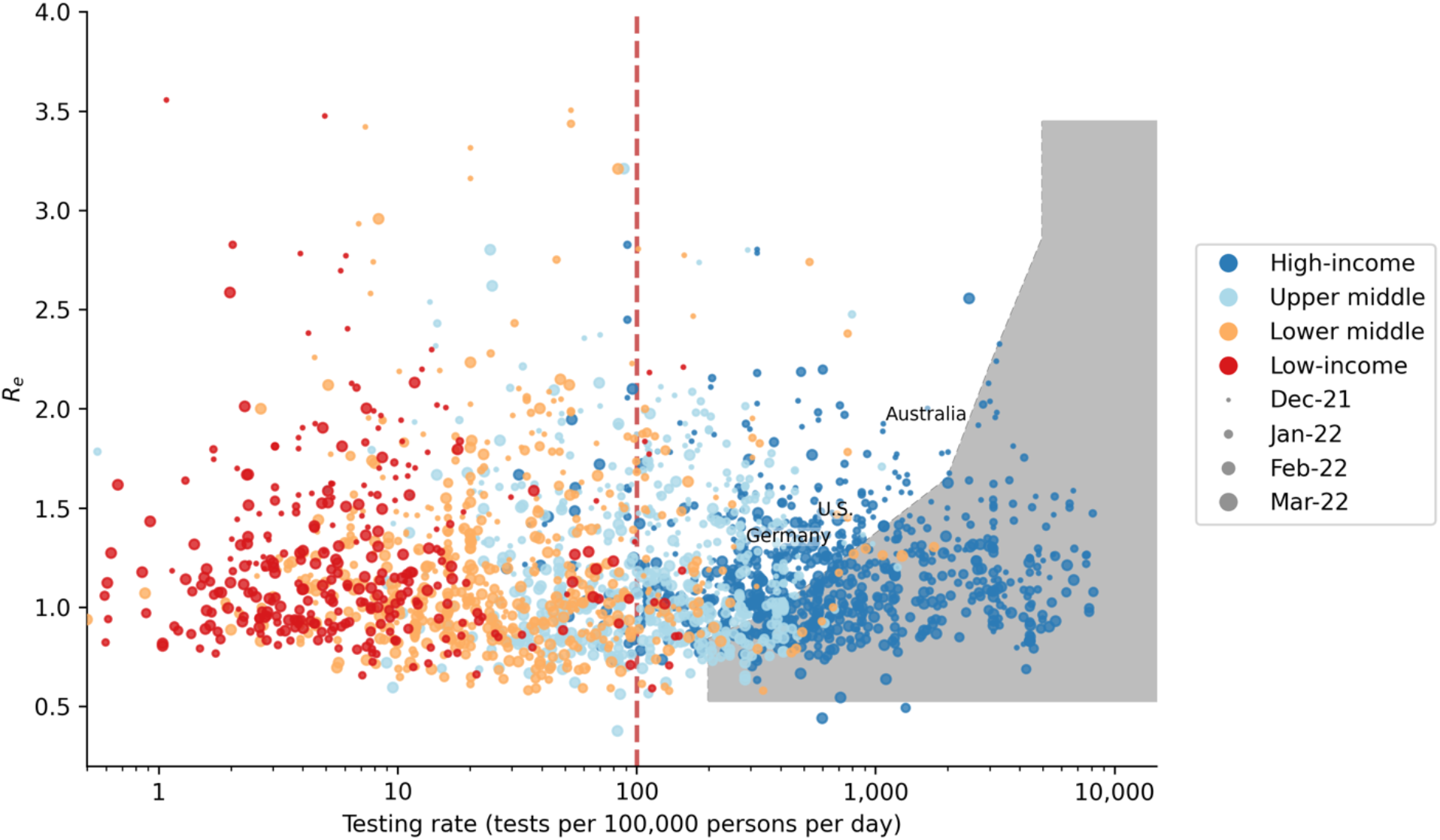
Global reported COVID-19 testing rate between December 2021 and March 2022 when the Omicron B.1 variant of concern spread rapidly across multiple countries. Each data point denotes the average weekly reported COVID-19 testing rate of a country against the average time-varying reproduction number (*R*_*t*_) computed in the same week and is colored by the income level of the country while sized by time (i.e. month/year). The shaded area denotes the level of test availability we had estimated to saturate symptomatic testing demand given different equivalent initial *R*_*e*_ values (Figure 4). The red vertical line at 100 tests per 100,000 persons per day is the minimum testing rate target set by the ACT-Accelerator diagnostics pillar. Testing rate data were sourced from the SARS-CoV-2 Test Tracker by FIND (https://www.finddx.org/covid-19/test-tracker/) while *R*_*t*_ was computed from reported COVID-19 case counts using EpiNow2 (https://github.com/epiforecasts/covid-rt-estimates).[28]

If there are excess tests available after meeting symptomatic testing demand, it is important to critically consider where and how routine community testing of asymptomatic populations is implemented to maximize impact. Given that a larger proportion of infections is expected to occur within households, household community testing after meeting symptomatic testing demand in the previous week would yield greater total infections averted. While testing at regular mass gatherings such as religious gatherings every week, for instance, could lead to comparable number of diagnosed infections, doing so only effectively tallies the number of infections that had happened in the week prior and limited infections at these gatherings. Disseminating tests across households, on the other hand, is more effective in not just lowering transmissions occurring in households but likely lessens the number of transmissions between different contact networks as well.

There are limitations with our study. First, we assumed that all healthcare facilities will have access to all Ag-RDT stocks available each week. However, there could be disparities in stock allocation between different clinics such as prioritizing stock allocation for hospitals. Such disproportionate distributions could lead to uneven fulfilment of symptomatic testing demand and consequently affect levels of infections. Furthermore, we assumed that there was a sufficient number of qualified health workforce and implementation support available to implement the various testing strategies. The strained healthcare system, especially in more remote regions of the country, poses a major limiting factor in implementing a widespread testing program. While our key finding that >100 tests/100k/day is needed to saturate symptomatic testing demand before rolling out community testing programs is unlikely to change, randomized community trials in LMICs using Ag-RDTs for community testing can provide better impact estimates under realistic scenarios.

Second, we only modelled scenarios where test-and-isolation was the only public health intervention. Symptomatic testing demand would expectedly be lower if other non-pharmaceutical interventions (NPIs) were introduced, and thus potentially improve the utility of community testing at lower test availability. However, the impact of NPIs is confounded by temporal effects[32] and thus difficult to parameterize their mean effects on infection control and in turn, testing demand. Since NPIs effectively decrease the number of secondary transmissions and in turn, *R*_*e*_, we expect that the testing demand for a population subjected to NPIs and testing would mirror the demand we had estimated for a population subject to testing only but at lower *R*_*e*_ values. Analogously, we also did not model how vaccination- and infection-acquired immunity affect testing demand explicitly. However, by the same reasoning that increased population immunity lowers *R*_*e*_, the testing demand for a partially immune population should be similar to that of a naïve population at lower *R*_*e*_ values as well.

Third, we parameterized incubation and virus shedding periods using those empirically measured from infections by wild-type (Wuhan-like) SARS-CoV-2[17,33] for this work. However, generation intervals have shortened considerably for recent VOCs such as Delta[34] and Omicron BA.1[35] and could impact the utility of testing in identifying an infection before it becomes infectious. We thus repeated the symptomatic-testing only simulations using incubation and virus shedding periods estimated for Omicron BA.1. There is effectively no difference in the amount of infections averted between the wild-type and Omicron BA.1 variant across all testing rates at *R*_*e*_ ≥ 1.5 (i.e. the expected initial effective reproduction number of the Omicron variant; Figure S5 and Supplementary Data).

To conclude, Ag-RDTs are a valuable diagnostic tool for COVID-19 testing capacities in LMICs. The target on minimal testing rate of 100 tests/100k/day should be seen as a true minimum if testing is going to be used for reducing transmission but substantially higher testing rates are needed to fulfil likely symptomatic testing demand or effective implementation of community testing.

## Data Availability

All data relevant to the study are included in the Article, the Supplementary Appendix and the GitHub repository (https://github.com/AMC-LAEB/PATAT-sim). The PATAT model source code can also be found in the GitHub repository
(https://github.com/AMC-LAEB/PATAT-sim).

## Data availability

All data relevant to the study are included in the Article, the Supplementary Data and the GitHub repository (https://github.com/AMC-LAEB/PATAT-sim). The PATAT model source code can also be found in the GitHub repository (https://github.com/AMC-LAEB/PATAT-sim).

## Acknowledgements

The authors are pleased to acknowledge that all computational work reported in this paper was performed on the Shared Computing Cluster which is administered by Boston University’s Research Computing Services (www.bu.edu/tech/support/research/).

## Authors’ contributions

A.X.H. contributed to the conceptualization, data curation, formal analysis, investigation, methodology, software, validation and visualization of the study. B.E.N. and C.A.R. contributed to the conceptualization, data curation, funding acquisition, investigation, methodology, project administration, resources, validation and supervision of the study. J.A.S., A.T., N.H. and E.H. contributed to the conceptualization, investigation, validation and visualization of the study. S.G. and S.K. contributed to the investigation, validation and visualization of the study. A.X.H. and C.A.R. wrote the original draft of the manuscript. All authors are involved in the review and editing of the manuscript. All authors had full access to all data of the study and the final responsibility for the decision to submit for publication.

