## Supplementary Data for "Strategies for using antigen rapid diagnostic tests to reduce transmission of SARS-CoV-2 in low- and middle-income countries: a mathematical modelling study applied to Zambia"

**Supplementary Data to Han et al. (2022)**

### Technical details of the Propelling Action for Testing and Treating (PATAT) simulation model

PATAT is a stochastic agent-based model designed to investigate the use and impact of antigen-detecting rapid diagnostic tests (Ag-RDT) in controlling COVID-19 outbreaks in low-middle income countries. The computational flow of a PATAT simulation is summarised as follows: First, an age-structured population of agents is created. Close contact networks are subsequently created based on the given demographic data. The simulation is then initialised and iterates over a given period of time where each time step corresponds to a day. The operations during each timestep encompass updating the disease progression of infected individuals, the status of isolated/quarantined agents, application of community testing strategies and computation of transmission events within contact networks.

#### Population demography

Using input demographic data which includes information such as population age and sex distribution, household composition, employment and schooling rates, PATAT generates a population of individuals who are linked by a series of underlying contact network settings where transmission may occur. These contact network settings include households, schools, workplaces, regular mass gatherings (i.e. church) as well as random community contacts.

#### Household

PATAT randomly generates a Poisson distribution of household sizes based on the given mean household size. A reference individual (e.g. head of the household) above an assumed prime adult age (e.g. years) is first randomly assigned to each household. To account for multigenerational households, the remaining household members are then randomly sampled multinomially by the input age distribution of households. Although PATAT does not explicitly model the geolocation of agents, households are ordered to implicitly approximate neighbourhood proximity.

#### Schools

PATAT distinguishes between elementary and secondary schools. For each education level, schooling children are randomly sampled from the population based on given enrolment rates and gender parity. Class sizes are then randomly drawn from a Poisson distribution based on the input mean class size while constrained by the number of schooling children attending the same grade (i.e. age; a class include only students studying the same grade). Schools are created by random allotment of classes such that (1) all schools will have equitable distributions of classes of all grades for the given education level and (2) the total number of students approximately equals to the expected school size. Classes are then populated by schooling agents such that (1) agents of proximally ordered households will tend to attend the same school and (2) children of the same grade (age) from identical households will not be assigned to the same class even though they may attend the same school. School teachers are then randomly drawn from the employed prime adult population based on the input teacher-to-student ratio and are assumed to have contact with each other during school days. Each class is randomly assigned to one teacher.

#### Workplaces

PATAT generates both formal and informal workplace contact networks based on separate employment rates. Youth (15-19 years) employment is also considered in the potential workforce. The distinction between formal and informal settings is made as mean employee contact rates likely differ between them. Furthermore, workplace distribution of Ag-RDTs for community testing is assumed to be feasible for formal employment entities only. Unlike schools, PATAT does not explicitly model for workplaces but sets up contact matrices between employed individuals who would be in regular contact at work. As such, different number of formal and informal mean number of work contacts must be provided by the user and sizes of workplace contact network are randomly drawn from a Poisson distribution. An employed agent would only be associated with one workplace contact network.

#### Mass gatherings (Churches)

High-density mass gatherings are considered in the model in the form of contacts among church congregations. The size of a church is assumed to follow a Normal distribution with user’s given mean and variance. PATAT assumes that all members of a household will visit a church together every Sunday. Other than close contacts with each other, each household member would also have a random number of close contacts from other households that attend the same church. This random contact number is drawn from a Gamma distribution with user’s given shape and scale parameters. Churches are also ordered such that proximally ordered households in the same neighbourhood would visit the same church.

#### Random community

PATAT assumes that every agent within a given age range would have a random number of contacts with the community daily, drawn from a Poisson distribution with a mean defined by the user.

#### Disease progression

PATAT implements a SEIRD epidemic model where the simulated population is distinguished between five compartments: susceptible, exposed (i.e. infected but is not infectious yet; latent phase), infected (which include the presymptomatic infectious period for symptomatic agents), recovered and dead. The infected compartments are further stratified by their presented symptoms, including asymptomatic, presymptomatic, symptomatic mild or severe. All symptomatic agents will also first undergo an infectious presymptomatic period after the exposed latent period. They will either develop mild symptoms who will always recover from the disease or experience severe infection which could either lead to death or recovery. PATAT uses age-structured wild-type SARS-CoV-2 disease severity and mortality probabilities that were also used in Covasim[1] (Table S1). As a simplification, PATAT currently assumes that all agents presenting severe symptoms will be hospitalized and removed from the population.

The total duration of infection since exposure depends on the symptoms presented by the patient and is comprised of different phases (i.e. latent, asymptomatic, presymptomatic, onset-to-recovery/death). The time period of each phase is drawn from the same distributions used by Covasim as well (Table S1).

#### Within-host viral dynamics

For each infected agent, PATAT explicitly simulates their viral load trajectory of cycle threshold (Ct) values over the course of their infection using a stochastic model modified from the one previously developed by Quilty et al.[2] A baseline Ct value ($Ct_{baseline}$) of 40 is established upon exposure. The infected agent becomes infectious upon the end of the latent period and their Ct value is assumed to be $\leq30$. A peak Ct value is then randomly drawn from a normal distribution of mean 22.3 and SD of 4.2.[3] Peak Ct is assumed to occur upon symptom onset for symptomatic agents and one day after the latent period for asymptomatic individuals. Cessation of viral shedding (i.e. return to $Ct_{baseline}$) occurs upon recovery or death. PATAT assumes that the transition rate towards peak Ct value should not be drastically different to that when returning to baseline upon cessation (i.e. there should be no sharp increase to baseline Ct value after gradual decrease to peak Ct value or vice versa). As such, the time periods of the different phases of infection are randomly drawn from the same quintile of their respective sample distribution. The viral load trajectory is then simulated by fitting a cubic Hermite spline to the generated exposed ($t_{exposed}$, $Ct_{baseline}$), latent ($t_{latent}$, $Ct_{latent}=30$), peak ($t_{peak}$, $Ct_{peak}$) and cessation values ($t_{recovered/death}$, $Ct_{baseline}$). The slope of the fitted curve is assumed to be zero for all of them except during $t_{latent}$where its slope is assumed to be $\frac{Ct_{peak}-Ct_{baseline}}{t_{peak}-t_{exposed}}$. PATAT then uses the fitted trajectory to linearly interpolate the viral load transmissibility factor ($f_{load,i}$) of an infectious agent $i$ assuming that they are twice as transmissible at peak Ct value (i.e. $f_{load}=2$) relative to when they first become infectious (i.e. Ct value = 30; $f_{load}=1$).

#### Transmissions

When an infectious agent $i$ comes into contact with a susceptible individual $j$, the probability of transmission ($p_{transmission, (i,j)}$) is given by:

$$p_{transmission,(i,j)}=\beta\times\Phi_{i}\times f_{c}\times f_{asymp,i}\times f_{load,i}\times f_{immunity,j}\times f_{susceptiblity,j}\times\rho_{i}\times\rho_{j}$$

where $\beta$ is the base transmission probability per contact, $\Phi_{i}$ is the overdispersion factor modelling individual-level variation in secondary transmissions (i.e. superspreading events), $f_{c}$ is a relative weight adjusting $\beta$ for the network setting $c$ where the contact has occurred, $f_{asymp,i}$ is the assumed relative transmissibility factor if infector $i$ is asymptomatic, $f_{immunity,j}$ measures the immunity level of susceptible $j$ against the transmitted virus (i.e. $f_{immunity,j}=1$ if completely naïve; $f_{immunity,j}=0$ if fully protected), $f_{susceptiblity,j}$ is the age-dependent susceptibility of $j$, $\rho_{i}$ and $\rho_{j}$ are the contact rates of infector $i$ and susceptible $j$ respectively.

$\Phi_{i}$is randomly drawn from a negative binomial distribution with mean of 1.0 and shape parameter of 0.45.[4] As evidence have been mixed as to whether asymptomatic agents are less transmissible, we conservatively assume there is no difference relative to symptomatic patients (i.e. $f_{asymp,i}=1$). The age-structured relative susceptibility values $f_{susceptiblity,j}$ are derived from odds ratios reported by Zhang et al.[5] (Table S1).

$\beta$ is determined by running initial test simulations with a range of values on a naïve population with no interventions that would satisfy the target basic reproduction number $R_{0}$ as computed from the resulting exponential growth rate and distribution of generation intervals.[6] $f_{c}$ is similarly calibrated during these test runs such that the transmission probabilities in households, workplaces, schools, and all other community contacts are constrained by a relative weighting of 10:2:2:1.[1]

#### Testing by Ag-RDT

Unlike PCR which is highly sensitive due to prior amplification of viral genetic materials, the sensitivity of Ag-RDT depends on the viral load of the tested patient. While the specificity of Ag-RDT is assumed to be 98.9%, its sensitivity depends on the Ct values of the tested infected agent: Ct $>35$ (0%); 35 – 30 (20.9%); 29 – 25 (50.7%); Ct $\leq24$ (95.8%).[7]

Testing by Ag-RDT may either occur via symptomatic testing at healthcare facilities or healthcare provided community testing. First, a symptomatic agent may opt to go into self-isolation upon symptom onset prior to being tested, as decided by a Bernoulli trial with probability $p_{self-isolation}$. Regardless if they were self-isolated, after $\tau_{delay,symp-test}$ days from symptom onset, the symptomatic agent may then decide to get tested with a Bernoulli probability of $p_{symp-test}$ that inversely correlates with the distance between the agent’s household and the nearest HCF (Table S1). PATAT assumes that agents who have decided against symptomatic testing (i.e. failed Bernoulli trial) or received negative test results will not seek symptomatic testing again.

For community testing in schools, given that teachers may act as inter-connecting agents linking between various classes, any available Ag-RDTs will always first be distributed to teachers in a school before they are distributed to students.

#### Isolation and quarantine

We assumed that agents would change their behaviour when (1) they start to present symptoms and go into self-isolation (10% compliance assumed, 71% endpoint adherence)[8]; (2) they test positive and are isolated for 10 days (50% compliance assumed, 86% endpoint adherence)[8]; or (3) they are household members (without symptoms) of positively-tested agents and are required to be in quarantine for 14 days (50% compliance assumed, 28% endpoint adherence)[8]. Once an agent goes into isolation/quarantine, we linearly interpolate their probability of adherence to stay in isolation/quarantine over the respective period. Given the lack of infrastructure and resources to set up dedicated isolation/quarantine facilities in many low-middle income countries, we assumed that all isolated and quarantined individuals would do so at home. Although they have no contact with agents outside of their home, we assumed that they would maintain 90% contact rate with household members.

#### Model Validation

To validate our model, we compared our simulation results against actual reported cases and deaths in Lusaka, Zambia between 25 December 2020 and 24 March 2021. Zambia was experiencing a second wave of infections as a result of the Beta variant.[9] Actual confirmed case and death tallies were retrieved from the Zambia COVID-19 Dashboard (<https://www.arcgis.com/apps/dashboards/3b3a01c1d8444932ba075fb44b119b63>). During this time, Zambia was performing ~40 tests/100,000 people/day[10]. We assumed that initial $R_{e}$ ~ 2.0 and simulated a 90-day epidemic wave under the aforementioned testing rate for 1,000,000 individuals using the demography parameters for Zambia (Table S1) and performed 10 independent simulations using PATAT. We multiplied the estimated mean number of reported (i.e. diagnosed) cases and deaths from our simulations by three to proportionally scale up the results for three million people, the approximate population size in Lusaka, Zambia. Our simulation results fit well against both actual reported case and death counts (Mean absolute difference = ~290 (case counts), 8 (deaths); Figure S6).

#### Sensitivity analysis with Omicron BA.1

We repeated the symptomatic-testing only simulations for a subset of $R_{e}$ values between 1.1 and 2.0 using incubation and virus shedding periods estimated for Omicron BA.1[11] (Figure S5). At $R_{e}=$ 1.1-1.2, we estimated that the shorter generational interval of Omicron would half the maximum proportion of infections averted upon saturating symptomatic testing demand (i.e. 20% for Omicron BA.1 as opposed to 40% infections averted for wild-type SARS-CoV-2) and a far greater number of tests would be needed to saturate symptomatic testing demand (i.e. 800-1000 tests/100k/day for Omicron BA.1 as opposed to 200-400 tests/100k/day for wild-type SARS-CoV-2). However, when $R_{e}\geq$ 1.5, the expected range of the initial $R_{e}$ of the Omicron BA.1 infection wave, there is effectively no difference in infections averted between the wild-type and Omicron variant across all testing rates.

### Supplemental Figures

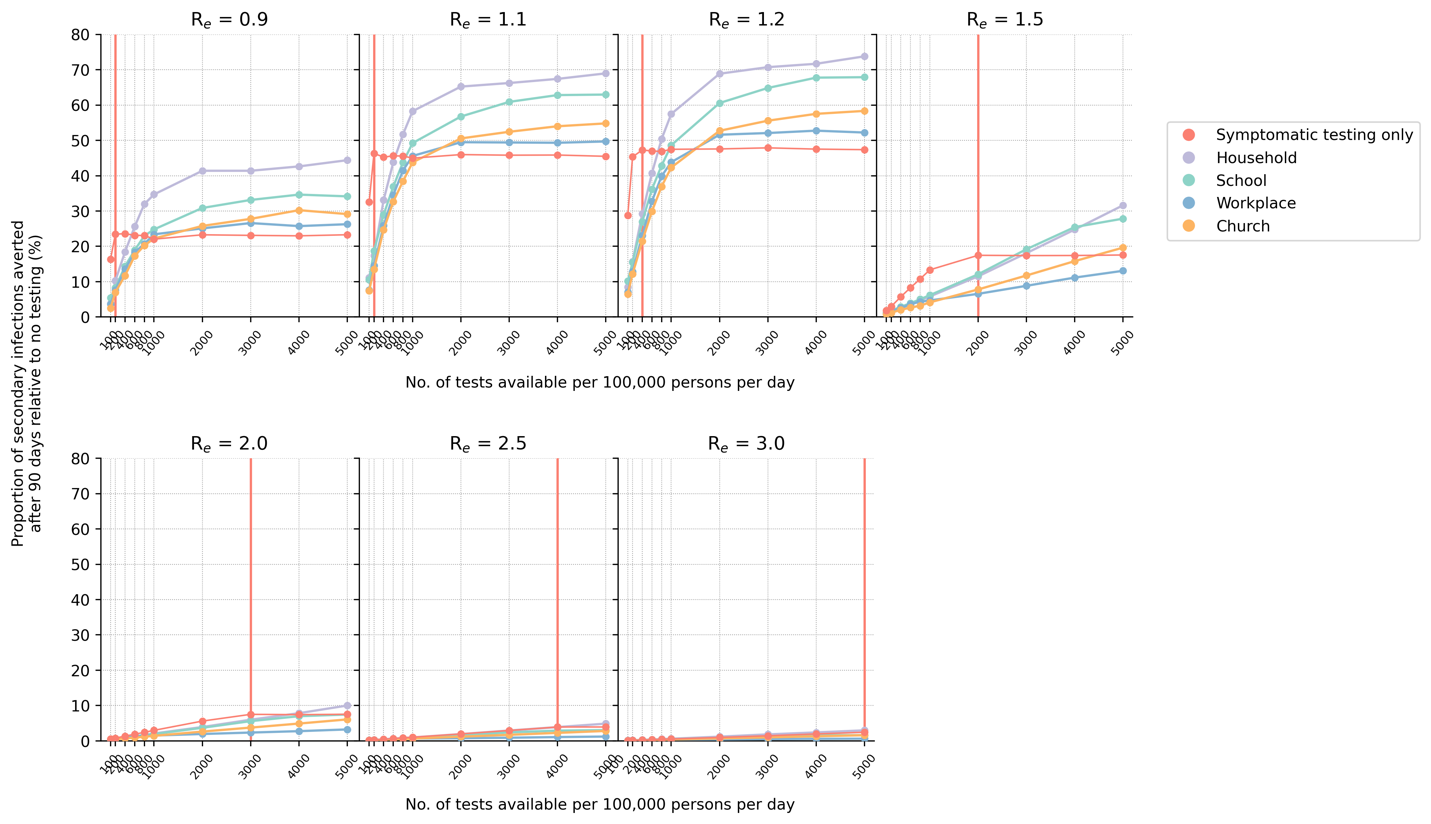

**Figure S1**: **Impact of either using all available Ag-RDT for symptomatic testing or a majority of them (85%) for community testing in various settings (even distribution only; with quarantine of household members)**. The proportion of secondary infections averted after 90 days relative to the no testing baseline for different number of tests available per 100,000 persons per day and assumed $R_{e}$ value is plotted for each test distribution strategy. The vertical red line denotes the number of tests required to saturate symptomatic testing demand.

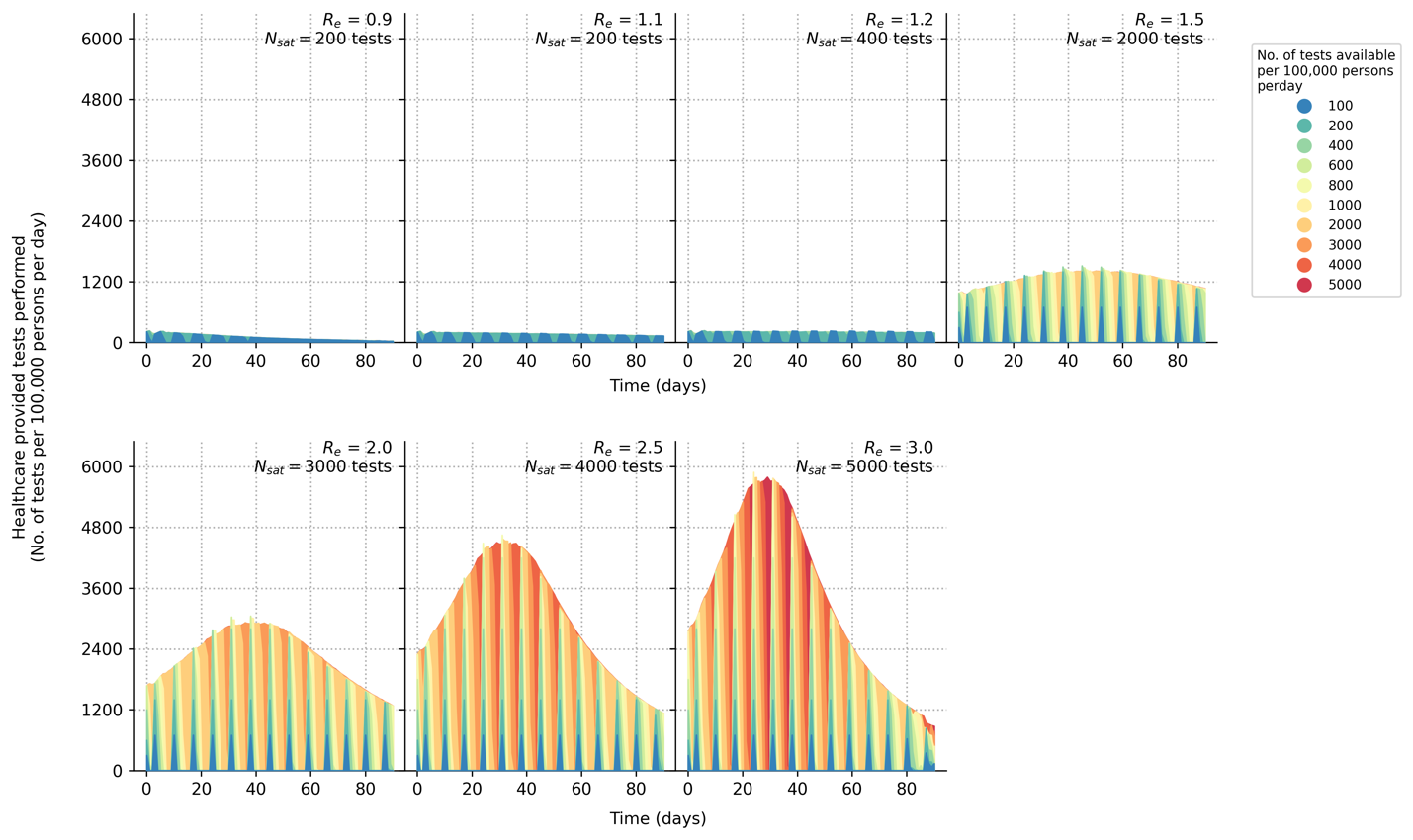

**Figure S2**: **Symptomatic testing demand during an epidemic (with quarantine of household members)**. Number of symptomatic tests performed per 100,000 persons per day over time for different initial $R_{e}$. Each differently colored shaded curve denotes a different number of tests available per 100,000 persons per day. We assumed that all healthcare facilities in the community will have new stocks of one week’s worth of Ag-RDTs every Monday. The symptomatic testing demand include both symptomatic SARS-CoV-2 infected agents who seek testing at healthcare facilities and those who seek symptomatic testing for other reasons based on assumed case positivity rates (see Methods). The area between the curve plotting number of tests needed to saturate symptomatic testing demand ($N_{sat}$) and any other curves plotting $N<N_{sat}$ is the amount of symptomatic testing shortage accumulated over time.

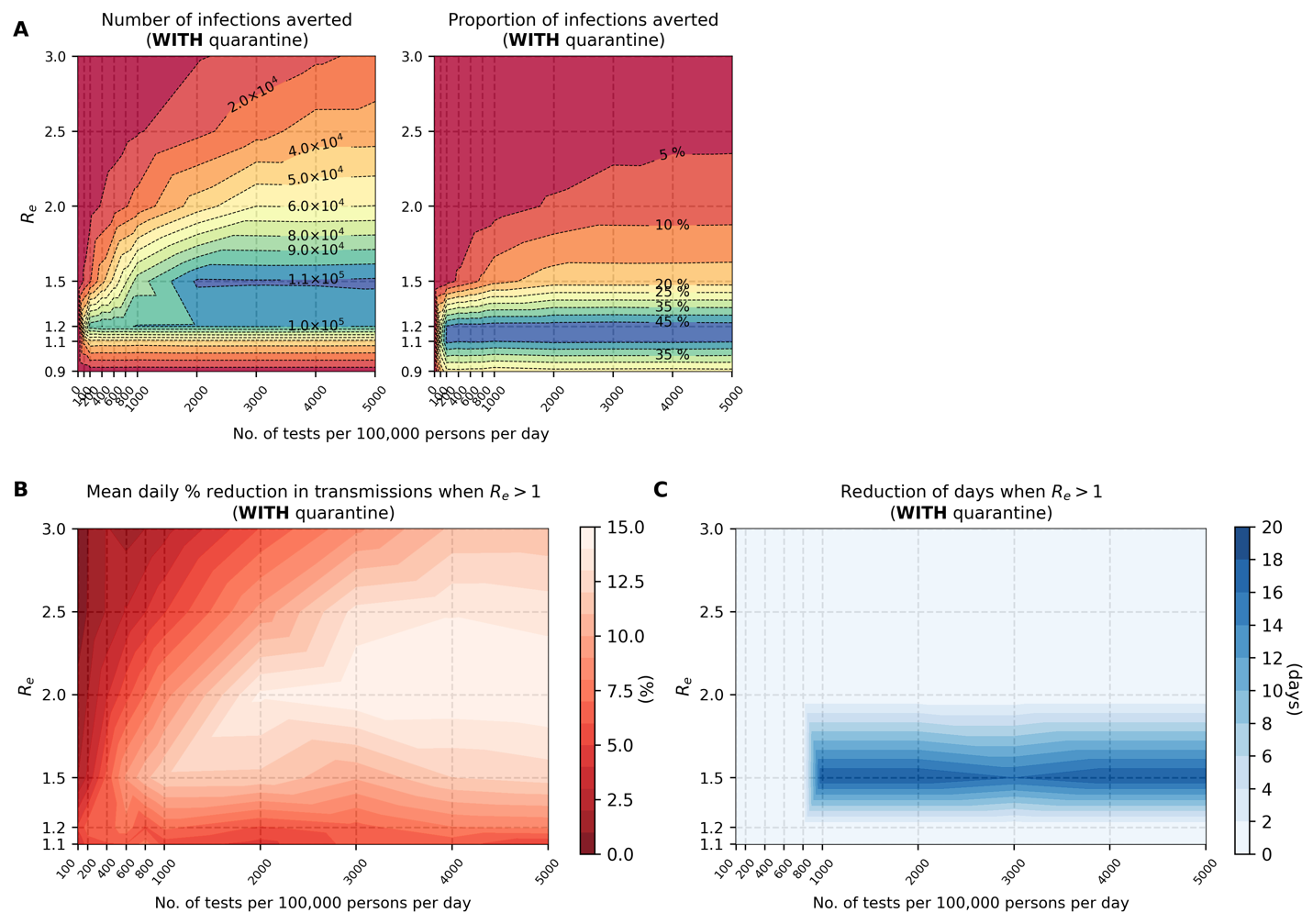

**Figure S3**: **Marginal impact of symptomatic testing prior to saturating demand (with quarantine of household members).** (**A**) Contour plots depicting infections averted relative to the no testing baseline for simulations with different initial $R_{e}$values and varying number of Ag-RDTs availability. Number of infections averted relative to no testing baseline after 90 days (left panel); Proportion of secondary infections averted relative to no testing baseline after 90 days (right panel). (**B**) Mean daily percentage reduction in transmissions while time-varying $R_{e}$ of simulated epidemic is still > 1 for different initial $R_{e}$values and varying number of Ag-RDTs available for symptomatic testing only. (**C**) Shortening of the number of days when time-varying $R_{e}$ of simulated epidemic is still > 1 for different initial $R_{e}$values and varying number of Ag-RDTs available for symptomatic testing only.

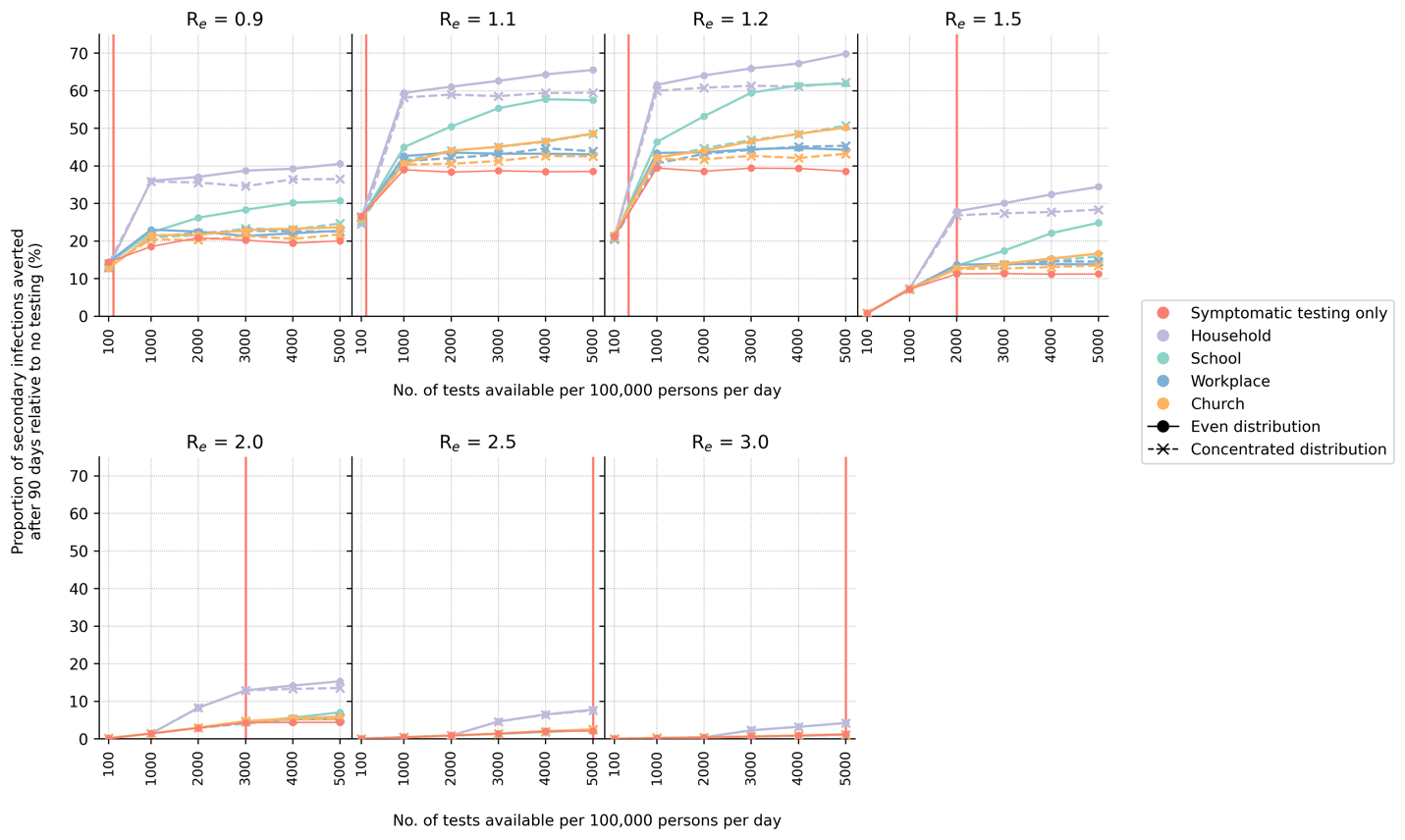

**Figure S4**: **Symptomatic-testing-first strategy to community testing (with quarantine of household members)**. When community testing is performed under this strategy, the leftover tests from the previous week’s stock allocated for symptomatic testing are used for community testing in various setting in the current week. Two different types of community test distributions approaches (even or concentrated; see Methods) were simulated. The proportion of secondary infections averted after 90 days relative to the no testing baseline for different number of tests available per 100,000 persons per day and assumed $R_{e}$ value is plotted for each test distribution strategy. The vertical red line denotes the number of tests required to saturate symptomatic testing demand.

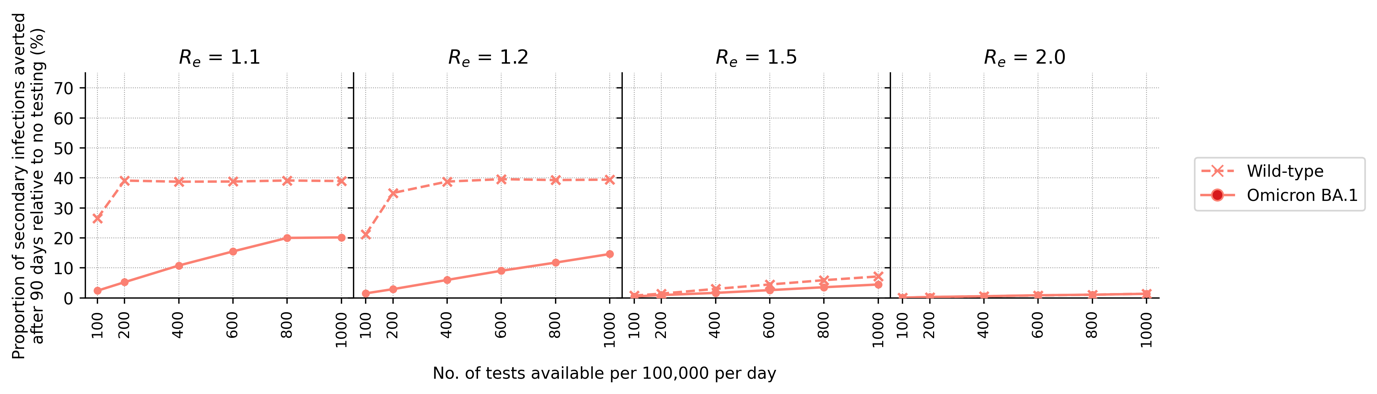

**Figure S5**: **Comparing impact of symptomatic testing only in an Omicron BA.1 wave against that for the wild-type (Wuhan-like) SARS-CoV-2 wave**. The proportion of secondary infections averted after 90 days relative to the no testing baseline for different number of tests available per 100,000 persons per day and assumed $R_{e}$ value is plotted.

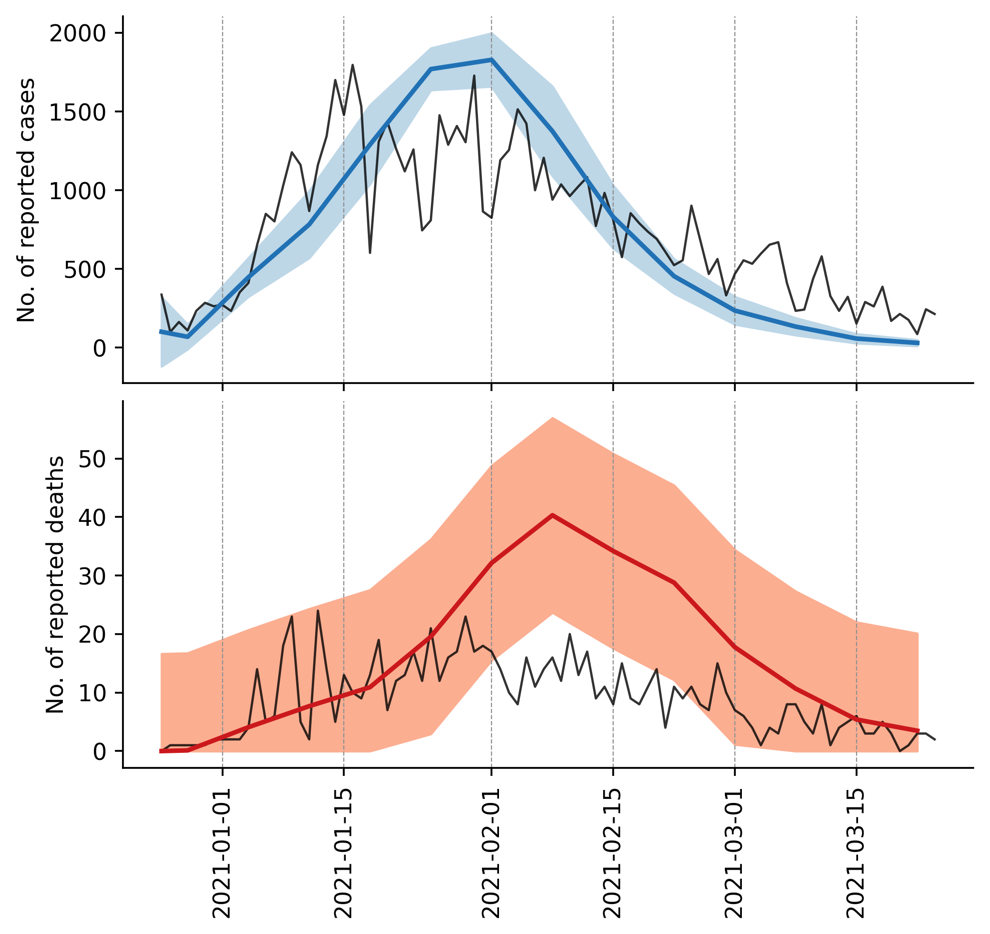

**Figure S6**: **Model validation**. We compared the mean number of reported cases (blue line, top panel) and deaths (red line, bottom panel) estimated by our simulations (10 simulations in total) against the actual case and death counts (black lines) in Lusaka, Zambia during the second wave of infections between 25 December 2020 and 24 March 2021. Actual case and death counts were retrieved from the Zambia COVID-19 Dashboard (<https://www.arcgis.com/apps/dashboards/3b3a01c1d8444932ba075fb44b119b63>). The blue and red shaded regions in each plot denotes the standard deviation of reported cases (top panel) and deaths (bottom panel) respectively.

### Supplemental Tables

**Table S1**: **PATAT simulation parameters**

| **Parameter** | **Values/Distribution** | **Source** |
| --- | --- | --- |
| *Population demography* | | |
| Total population size | 1,000,000 |  |
| Mean household size | 5.0 | [12] |
| Age structure (in bins of 5 years) | [0.161, 0.165, 0.157, 0.101, 0.083, 0.068, 0.057, 0.051, 0.042, 0.030, 0.024, 0.015, 0.016, 0.009, 0.008, 0.005, 0.006, 0.002, 0.000, 0.000] | [12] |
| Minimum prime adult age | 20 years | Assumed |
| Proportion of women | 51% | [13] |
| Minimum working age | 15 years | [13] |
| Employment rate | 39% (male), 23% (female) | [13] |
| Formal employment rate | 36% (employed male), 24% (employed female) | [13] |
| Schooling rate | 79% (male), 40% (female) | [12] |
| School gender parity | 1.0 (Primary), 0.9 (Secondary) | [12] |
| Church participation rate | 70% of all households | Assumed |
| Mean employment contacts (formal) | 20 | Assumed |
| Mean employment contacts (informal) | 5 | Assumed |
| Mean class size | 37 (Primary and secondary) | [12] |
| Mean school size | 700 (Primary and secondary) | Assumed |
| Student-teacher ratio | 42 (Primary and secondary) | [12] |
| Mean church size (s.d.) | 500 (100) | Assumed |
| Mean random contacts in church per person | 10 | Assumed |
| Mean random community contacts per day | 10 | Assumed |
| *SARS-CoV-2 transmissions related parameters* | | |
| Age-structured relative susceptibility (in bins of 5 years) | [0.34, 0.34, 0.67, 0.67, 1.00, 1.00, 1.00, 1.00, 1.00, 1.00, 1.00, 1.00, 1.00, 1.00, 1.24, 1.24, 1.47, 1.47, 1.47, 1.47] | [1,5] |
| Age-structured probability of becoming symptomatic (in bins of 5 years) | [0.50, 0.50, 0.55, 0.55, 0.60, 0.60, 0.65, 0.65, 0.70, 0.70, 0.75, 0.75, 0.80, 0.80, 0.85, 0.85, 0.90, 0.90, 0.90, 0.90] | [14,15] |
| Age-structured probability of developing severe disease (in bins of 5 years) | [0.00050, 0.00050, 0.00165, 0.00165, 0.00720, 0.00720, 0.02080, 0.02080, 0.03430, 0.03430, 0.07650, 0.07650, 0.13280, 0.13280, 0.20655, 0.20655, 0.24570, 0.24570, 0.24570, 0.24570] | [14,15] |
| Age-structured probability of death (in bins of 5 years) | [0.00002, 0.00002, 0.00002, 0.00002, 0.00010, 0.00010, 0.00032, 0.00032, 0.00098, 0.00098, 0.00265, 0.00265, 0.00766, 0.00766, 0.02439, 0.02439, 0.08292, 0.08292, 0.16190, 0.16190] | [16,17] |
| Latent period (days) | Lognormal (4.5, 1.5) | [1,18] |
| Pre-symptomatic period (days) | Lognormal (1.1, 0.9) | [1,18] |
| Period between symptom onset and severe disease (days) | Lognormal (6.6, 4.9) | [18] |
| Period between severe disease and death (days) | Lognormal (8.6, 6.7) | [18] |
| Recovery period for symptomatic agents with mild disease (days) | Lognormal (8.0, 2.0) | [19] |
| Recovery period for asymptomatic agent (days) | Lognormal (8.0, 2.0) | [19] |
| Recovery period of agents with severe disease (days) | Lognormal (18.1, 6.3) | [14] |
| *Testing parameters* | | |
| Delay in visiting healthcare facility for symptomatic testing (days) | Lognormal (1.0, 0.5) | Assumed |
| Ag-RDT specificity | 0.989 | [7] |
| Agents to healthcare facilities ratio | 7,000:1 | [20,21] |
| Distance-structured distribution of households to nearest healthcare facility (in bins of 1km) | [0.048, 0.193, 0.119, 0.08, 0.074, 0.098, 0.068, 0.072, 0.056, 0.191] | [22] |
| Distance-structured probabilities of agent visiting nearest healthcare facility for symptomatic testing (in bins of 1km) | [0.853, 0.808, 0.762, 0.717, 0.672, 0.626, 0.581, 0.536, 0.49, 0.445] | [22] |
| *Isolation/quarantine parameters* | | |
| Isolation period | 10 days |  |
| Quarantine period | 14 days |  |
| Self-isolation period | 10 days |  |
| Reduction in contact rates under isolation/quarantine (in order of households, schools, workplaces, church and random community) | [10%, 100%, 100%, 100%, 100%] |  |
